# Longitudinal Analysis of Electronic Health Records Reveals Medical Conditions Associated with Subsequent Alzheimer’s Disease Development

**DOI:** 10.1101/2025.03.22.25324197

**Authors:** Xue Zhong, Gengjie Jia, Zhijun Yin, Kerou Cheng, Andrey Rzhetsky, Bingshan Li, Nancy J. Cox

## Abstract

**Background:** Several health conditions are known to increase the risk of Alzheimer’s disease (AD). We aim to systematically identify medical conditions that are associated with subsequent development of AD by leveraging the growing resources of electronic health records (EHRs).

**Methods:** This retrospective cohort study used de-identified EHRs from two independent databases (MarketScan and VUMC) with 153 million individuals to identify AD cases and age– and gender-matched controls. By tracking their EHRs over a 10-year window before AD diagnosis and comparing the EHRs between AD cases and controls, we identified medical conditions that occur more likely in those who later develop AD. We further assessed the genetic underpinnings of these conditions in relation to AD genetics using data from two large-scale biobanks (BioVU and UK Biobank, total N=450,000).

**Results:** We identified 43,508 AD cases and 419,455 matched controls in MarketScan, and 1,320 AD cases and 12,720 matched controls in VUMC. We detected 406 and 102 medical phenotypes that are significantly enriched among the future AD cases in MarketScan and VUMC databases, respectively. In both EHR databases, mental disorders and neurological disorders emerged as the top two most enriched clinical categories. More than 70 medical phenotypes are replicated in both EHR databases, which are dominated by mental disorders (e.g., depression), neurological disorders (e.g., sleep orders), circulatory system disorders (e.g. cerebral atherosclerosis) and endocrine/metabolic disorders (e.g., type 2 diabetes). We identified 19 phenotypes that are either associated with individual risk variants of AD or a polygenic risk score of AD.

**Conclusions:** In this study, analysis of longitudinal EHRs from independent large-scale databases enables robust identification of health conditions associated with subsequent development of AD, highlighting potential opportunities of therapeutics and interventions to reduce AD risk.

## INTRODUCTION

Alzheimer’s disease (AD) poses a huge healthcare burden in the United States and globally. AD is an age-related neurodegenerative disorder characterized by an initial decline in memory and subsequent impairment of executive and cognitive functions^1^. Until recently, there were no effective disease-modifying treatments available for AD. Since 2021, three anti-amyloid drugs have been FDA-approved that demonstrate efficacy in removing the pathological amyloid proteins and slowing cognition decline in patients with early Alzheimer’s disease (mild cognitive impairment or mild dementia due to Alzheimer’s disease) ^2–4^. It is known that AD develops over decades, with the gradual accumulation of amyloid and tau proteins in the brain before the clinical manifestations and lapse in memory and cognition. Several health conditions have been linked to an increased risk of developing Alzheimer’s disease late in life. For example, midlife hypertension, hyperlipidemia, and stroke have been shown to increase the incidence of AD two decades later^5,6^; changes in vision among adults over the age of 50 have been associated with the accumulation of two hallmark proteins of AD in the brain^7,8^; and depression was more frequently observed in individuals who later received an AD diagnosis compared to their peers^9,10^. While the nature of these conditions in relation to AD may vary (some are risk factors, others may serve as early manifestations), they are all associated with a higher incidence of AD. These conditions may enable earlier recognition of functional changes associated with AD as well as potential intervention long before the hallmark clinical symptoms of memory and/or cognition impairment become apparent. It is projected that delaying the onset of AD by just five years could halve the incidence rate^11^.

The current inventory of medical conditions that predict the future development of AD is limited. Longitudinal electronic health records (EHRs) enable the tracking of disease progression and may reveal functional changes associated with AD before the onset of memory and/or cognition lapses. Numerous studies have established that diagnosis codes (billing codes, ICD codes) of AD within EHRs are sufficiently accurate for research purposes, with a positive predictive value of ∼75% for AD diagnosis codes in most health systems^12–14^. We propose that longitudinal EHRs from millions of individuals can facilitate a systematic identification for medical conditions that predict the future onset of AD.

Here, we systematically mined longitudinal EHR data in MarketScan, a national-level claim-based database with over 150 million individuals, as the discovery cohort, and identified medical phenotypes that are significantly enriched in those who later receive an AD diagnosis. We analyzed the longitudinal data in VUMC’s hospital-based EHR system with about 3 million patients as an independent replication cohort to validate the findings. also used an independent, large-scale EHR database for replication analysis. To further tease apart clinical phenotypes that are potential mediators of AD, we assessed the genetic basis of the robustly identified medical phenotypes through a phenome-wide association (PheWAS)^15^ analysis of genetic instruments constructed using the latest genome-wide association study (GWAS) of AD and related dementia^16^.

## METHODS & MATERIALS

The workflow and study design are illustrated in **Figure 1**.

**Figure 1.**
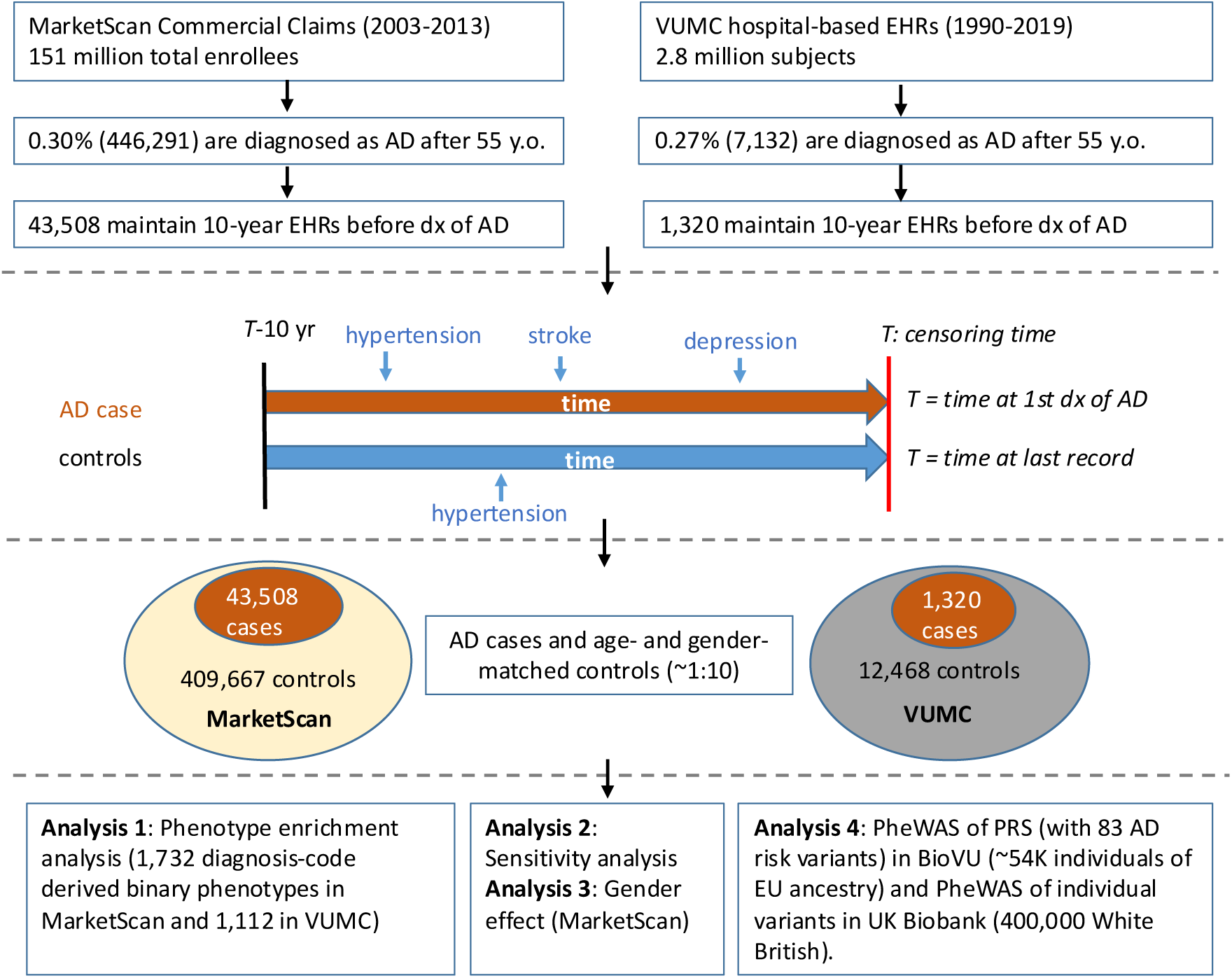
Study workflow and matching design. MarketScan: a commercial, claim-based national repository of EHRs; VUMC EHRs: a hospital-based, de-identified EHRs of 2.8 million unique patients from Vanderbilt University Medical Center; PheWAS: phenome-wide association study; PRS: polygenic risk score. Cases of Alzheimer’s disease are identified using diagnosis codes given by the international classification of disease (ICD) version 9 and version 10. Each AD case was matched up to 10 controls by age, gender and years of EHRs prior to their censoring time.

### Data sources

The MarketScan claims database contains US national-level collection of health records for over 151 million individuals enrolled between 2003 and 2013 ^17^. This database comprises insurance claim-based records that include inpatient and outpatient medical events, procedures, medications, and healthcare expenditures. The data were sourced from numerous private insurance companies, managed care organizations, health plan providers, and state Medicaid agencies. The covered patient population therefore represent affluent, privately insured segments of US society ^17,18^.

The Synthetic Derivative (SD) of Vanderbilt University Medical Center (VUMC) is a hospital-based EHR database, containing de-identified version of the entire EHRs of VUMC from about 3 million unique individuals. The SD includes diagnostic and procedure codes, hospital admissions, provider progress notes, discharge summaries, laboratory data, and medication histories, from which researchers can extract all phenotypes of interest, such as disease diagnoses and drug effects. The SD data are updated regularly, and for this study, we extracted all available data till the end of January 2020. The use of SD in research projects is classified as non-human subject research by the VUMC Institutional Review Board.

BioVU is the biobank of VUMC, which houses DNA samples from more than 250,000 individuals from the SD. Samples selected for genotyping are enriched for patients whose medical home is VUMC. A detailed description of program operations, ethical considerations, and continuing oversight and patient engagement has been published ^19^. Genotype data from BioVU were imputed using the Haplotype Reference Consortium reference panel (version r.1.1) ^20,21^ with the IMPUTE2 software ^22^. For analysis, samples of European ancestry were selected based on principal component analysis (PCA) of genetic ancestry.

### Identify AD cases and matched controls

Previous studies have shown that the diagnosis codes of AD recorded in EHRs achieve sufficient accuracy for research purposes, with a positive predictive value of 75% for AD diagnosis codes across most health systems ^12,14^. In this study, we used the International classification of disease codes (ICD) version 9 and version 10 (ICD9: 331.0, 3310.00; ICD10: F00, F00,0, F00.1, F00.2; ICD-10: G30, G30.0, G30.1, G30.8, G30.9) to identify cases of AD. We required a minimum of two codes on separate visit dates to instantiate an AD case. We defined controls as individuals without any AD diagnosis codes in their records up to their last visit. To enhance statistical power, we matched each AD case with four to ten controls (if more than 10 controls are available to match an AD case, we randomly sampled 10 control individuals to comprise the control set for this AD case; if less than 4 controls are available to match an AD case, we do not consider that AD case nor the controls). The matched controls were required to be of the same gender as the AD case and same age at the censoring time of the AD case (i.e., the date of the first AD diagnosis). Both AD cases and their matched controls were required to maintain at least 10 years of EHRs before the “index time.” The index time for an AD case is same as the censoring time; the index time of the corresponding controls matched to that AD case is the censoring time defined for that AD case.

### Medical phenotype definition

We used phenotypes derived from the billing codes of electronic health records, as previously described ^15^, to define medical phenotypes. The resulting binary phenotypes are referred to as ‘phecodes,’ each of which may correspond to multiple diagnosis codes (ICD9 or ICD10). For example, the phecode for *dementia with cerebral degenerations* encompasses a range of ICD codes, including frontotemporal dementia (ICD9 code 331.1) and dementia with Lewy bodies (ICD9 331.82). Each phecode has defined case, control and exclusion criteria, and we required a minimum of two codes on different visit dates to instantiate a case for each phecode.

### Statistical analysis

For each binary phenotype (‘phecode’), we compared the number of cases (unique individuals) between AD patients and matched controls over a 10-year preceding period. Fisher’s exact test (function *fisher.test* in R package “stats”) was employed to assess the relative enrichment of the associated medical event in AD patients compared to controls. We required each phecode has at least five cases in AD and non-AD groups to meet the criterion of applying the Fisher’s exact test. The analysis was performed in MarketScan and VUMC separately. At each site, we applied Bonferroni correction to determine the threshold for statistical significance. A total of 1,722 phecodes in MarketScan and 1,112 phecodes in VUMC were analyzed for enrichment. To identify disproportionately enriched clinical categories for AD-enriched phenotypes (phecodes), we compared the proportion of the enriched phecodes per clinical category to the overall proportion of enriched phecodes (i.e. overall proportion=466/1722=0.27 for MarketScan and overall proportion=102/1112=0.92 for VUMC) using the one-sided proportion test (function *prop.test* in R package “stats” with the parameter *alternative= “greater”*). Similarly, to identify the disproportionately depleted clinical categories for AD-depleted phenotypes, we compared the proportion of the depleted phecodes per clinical category to the overall proportion of depleted phecodes (i.e. overall_proportion=106/1722=0.06 for MarketScan and overall_prop=2/1112=0.0018 for VUMC).

### Sensitivity analysis

The co-occurrence of an AD diagnosis with diagnosis of other dementia types appeared common in EHRs. To assess how sensitive the enrichment estimation is affected due to such comorbidity, we constructed a sensitivity analysis in the larger database, MarketScan. Specifically, in MarketScan, we excluded AD cases who also carried diagnoses of other dementia types, including *vascular dementia*, *frontotemporal dementia*, *dementia with Lewy bodies*, *dementias*, *dementia with cerebral degeneration*, *delirium dementia and amnestic and other cognitive disorders*, and *senile dementia*. Correspondingly, we also removed the controls matched to such AD cases. This resulted in a subset with 13,391 AD cases and 133,910 matched controls involving 1644 phecodes feasible for the Fisher exact test, on which we repeated the enrichment analysis. Bonferroni correction threshold of 3.04×10^−5^ (=0.05/1644) was used to determine the significance of associations.

### Polygenic risk score

The genetic instruments we used to assess the relationship between the enriched phenotypes and AD genetics include risk variants of AD and an aggregate of these variants as a polygenic risk score. The polygenic risk score (PRS) of AD was constructed using the APOE locus and 83 independent signals as reported from the largest GWAS study of AD and related dementia ^16^. These 84 risk variants will also be assessed for variant-level phenome-wide associations The PRS is calculated as a weighted sum of the dosage of each AD-risk-increasing allele multiplied by the effect size (natural logarithm of odds ratio) of that allele. In BioVU genotyped data, there are 79 risk variants available for analysis (see **Suppl. Table S1**), which were used to calculate the PRS. The resulting polygenic risk score of BioVU individuals of European ancestry was used in PheWAS analysis.

### PheWAS

A systematic interrogation of phenotypic associations of genetic instrument variables of AD was performed in BioVU and UK Biobank using the PheWAS approach. In BioVU (n=54000 individuals of European ancestry), PheWAS of PRS and PRS excluding the APOE locus were performed. This was achieved through logistic regression, adjusted for age, gender, three principal components of ancestry, and arrays/batches, utilizing the PheWAS package ^23^. Only phecodes with at least 20 cases were included in analysis. In addition, PheWAS of the APOE locus (rs429538_C) was also performed in BioVU. The variant-level phenome-wide associations of a total of 84 AD risk variants (**Suppl. Table S1**) are based on the UK Biobank data and the Pheweb source (https://pheweb.sph.umich.edu) was used to extract the association results. The Pheweb displays GWAS results for ∼1400 binary phecodes (derived from EHR ICD billing codes) across 28 million variants that were genotyped and imputed using TOPMed ^24^ from ∼400,000 White British individuals of the UK Biobank. Analyses on binary outcomes were conducted using SAIGE ^25^ (a method of association test accounting for case-control imbalance that is typical of EHR data), adjusting for genetic relatedness, sex, birth year and the first 4 principal components.

## RESULTS

### Characteristics of study cohorts

Utilizing the MarketScan (MS) database, a commercial, claim-based national repository of EHRs encompassing over 150 million individuals from 2003 to 2013, we identified 446,291 individuals (∼0.3% of the population) with a diagnosis code for AD recorded after age 55. The majority (∼90%) of these AD diagnoses, approximated by the first appearance of the AD ICD code, occurred after 65 years old. From this group, 43,508 individuals (56% female) with at least 10 years of EHRs prior to their first AD diagnosis were selected. Each AD case was then paired with 4 to 10 controls of equivalent age and gender who also had at least 10 years of EHRs prior to the censoring date (i.e. the date of last follow-up) and did not have an AD diagnosis code. In total, we obtained 43,508 AD cases and 419,455 matched controls from the MS database (**Table 1**). Using the same methodology, we identified 1,320 AD cases and 12,720 age– and gender-matched controls from the de-identified EHRs of about 3 million individuals at VUMC as of Jan 2020 (**Table 1**). Notably, a consistent proportion of AD cases was observed across both databases (0.30% in MS and 0.27% in VUMC), predominantly among females (56% in MS and 62% in VUMC), with the age of diagnosis predominantly above 75 years (84% in MS and 81% in VUMC) (**Table1**).

**Table 1.** Subject characteristics and sample size from two EHR databases.

|  | MarketScan | VUMC |
| --- | --- | --- |
| Description | Insurance claims-based | Hospital-based |
| Sample size | 151 million | 2.8 million |
| Diagnosed (dx) with AD, *n* (female%) | 446,291 (female 56%) | 7,132 (female 63%) |
| Stratify by age, *n* (%) |  |  |
| <65 y | 52,579 (12%) | 613 (9%) |
| 65-74 y | 86,703 (19%) | 1411 (20%) |
| 75-84 y | 194,257 (44%) | 3469 (49%) |
| >=85 y | 112,752 (25%) | 1639 (23%) |
| >10y EHRs before 1<sup>st</sup> AD dx & 1<sup>st</sup> dx at 55 y.o. or later, *n* (female%) | 43,508 (?) | 1,320 (?) |
| Stratify by age at 1st dx, *n* (%) |  |  |
| 55-64 y | 1,859 (4%) | 17 (2%) |
| 65-74 y | 5,292 (12%) | 237 (18%) |
| 75-84 y | 17,431 (40%) | 698 (53%) |
| >=85 y | 18,926 (44%) | 368 (28%) |
| Sum | 43,508 (100%) | 1,320 (100%) |
| Matched Controls, *n* (female%) | 409,677 (?) | 12,468 (?) |

For the identified AD cases and controls, we subsequently traced their EHRs over a 10-year period prior to an index date. For the AD case, the index date corresponds to the date of their first AD diagnosis; whereas for the controls, it is the censoring date (*i.e.*, the date of their last recorded visit within the dataset). We compared the prevalence of each medical condition available in the records between the case and control groups to pinpoint conditions that occurred more often in those who later developed AD. In the two EHR databases, we employed phecodes (binary phenotypes derived from ICD billing codes)^15^ to delineate the “case” and “control” status for each respective medical phenotype.

### Discovery in MarketScan

In the MarketScan database, a total of 1,722 phecodes were tested for their association with future AD cases, and 406 phecodes were significantly enriched in future AD cases compared to the matched controls after Bonferroni correction (at a significance of *P*=0.05/1722=2.90×10^−5^). Conditions such as hypertension, hyperlipidemia, and type 2 diabetes (T2D) were more prevalent in those who later developed AD, aligning with previous observations based on prospective cohorts ^6^. Memory loss (OR=8.0, *P*<10^−321^) and mild cognitive impairment (OR=6.0, *P*<10^−321^), two canonical clinical manifestations of AD, were among the most enriched phenotypes. This was followed by other dementia types, such as *vascular dementia* (OR=6.4, *P*<10^−321^) and *dementia with cerebral degenerations* (OR=4.8, *P*<10^−321^). Other highly enriched phenotypes included a range of psychological symptoms, such as *paranoid disorders* (OR=6.3, *P*<10^−321^), *psychosis* (OR=4.6, *P*<10^−321^), *mood disorders* (OR=3.4, *P*<10^−321^), *depression* (OR=2.4, *P*<10^−321^), and *transient alteration of consciousness* (OR=2.3, *P*<10^−321^); neurological disorders such as *encephalopathy* (OR=3.1, *P*<10^−321^), *cerebral degeneration* (OR=2.6, *P*<10^−321^), *Parkinson’s disease* (OR=2.6, p<10^−321^), *lack of coordination* (OR=2.0, *P*<10^−321^), and *abnormality of gait* (OR=1.9, p<10^−321^); as well as circulatory system issues such as *cerebral ischemia* (OR=2.4, *P*<10^−321^), *cerebral artery occlusion with cerebral infarction* (OR=1.9, *P*<10^−321^), and *cerebrovascular disease* (OR=1.9, *P*=9.0×10^−172^) (see **Suppl. Table S2** for the full list).

Since each phenotype can be categorized within a broader clinical domain (totaling 17 categories, see **Suppl. Table S2**), we then investigated whether certain clinical categories disproportionately contributed to the enrichment of phenotypes associated with AD. After assigning the phecodes to their respective clinical categories, we assessed the enrichment of each category using Bonferroni correction (at a significance of *P*=0.05/17=0.0029), while considering the size of each category (for example, the *circulatory systems* category contains twice as many phenotypes as the *mental disorders* category, thereby potentially contributing to a greater number of enriched phenotypes). We found that *mental disorders*, a clinical category containing 63 phecodes, was among the most significantly enriched, with 56 of these phecodes showing enrichment (56/63 compared to the overall proportion 406/1722, proportion test *P*=8.2×10^−34^). Other top enriched clinical categories included *neurological disorders* (49/69, *P*=3.1×10^−20^), *injuries & poisonings* (60/87, *P*=3.5×10^−23^), *symptoms* (22/37, *P*=3.7×10^−7^) and *circulatory system* (53/128, *P*=1.7×10^6^) (**Suppl. Table S3**). Collectively, these clinical categories, which account for 22% of the medical phenotypes, contribute to 59% (240/406) of the phenotypic enrichment observed.

Finally, we reported a list of 166 phenotypes that are depleted in the pre-AD EHRs, with a quarter of such phenotypes coming from the *neoplasms* category (**Suppl. Table S4**).

### Replication in VUMC

We conducted the same analysis within the VUMC database. Out of the 1,112 phecodes examined, we detected 102 significantly enriched phenotypes (at the Bonferroni correction threshold, *P*=0.05/1112=4.5×10^−5^) (see **Suppl. Table S4**). In alignment with the findings from MarketScan, *essential hypertension* (OR=1.8, *P*=1.2×10^−23^), *hyperlipidemia* (OR=1.6, *P*=2.9×10^−18^), and *type 2 diabetes* (OR=1.3, *P*=1.3×10^−5^) are on the list; *memory loss* (OR=13.3, *P*=1.3×10^−318^), *vascular dementia* (OR=6.3, *P*=1.4×10^−44^), *depression* (OR=3.3, *P*=5.0×10^−65^), *anxiety disorder* (OR=2.7, *P*=1.2×10^−35^), *paranoid disorders* (OR=8.8, *P*=3.3×10^−25^), *cerebral degeneration* (OR=2.6, *P*=2.8×10^−13^), and *cerebral ischemia* (OR=2.1, *P*=1.6×10^−8^) were among the most highly enriched phenotypes. The clinical categories of *mental disorders* (26/52 compared to the overall proportion 102/1112, proportion test *P*=1.1×10^−23^) and *neurological disorders* (12/50, *P*=0.0004) also emerged as the top categories with disproportionate and significant enrichment (**Suppl. Table S3**). Notably, the only phenotypes that were significantly depleted were found within the *neoplasms* category (**Suppl. Table S5**).

### Robustly enriched phenotypes in both MarketScan and VUMC datasets

We have compiled a list of 73 phenotypes that are significantly enriched in both the MarketScan and VUMC EHR datasets after Bonferroni correction (i.e., *P*<2.9×10^−5^ in MS and *P*<4.5×10^−5^ in VU) (**Table 2**). These phenotypes, which span nine clinical categories, are predominantly represented by two: *mental disorders* (27 phecodes) and *neurological disorders* (12 phecodes). Specifically, speech disorders (*aphasia/speech disturbance*, *aphasia*, *speech and language disorder*) were more commonly seen in those who later developed AD. Severe psychological problems such as *paranoid disorders*, *psychosis*, *hallucinations* and *suicidal ideation* were also significantly enriched (all with OR>3) over the years prior to AD diagnosis. Neurological manifestations associated with the future development of AD included *abnormal involuntary movements, convulsions* (involuntary muscle contractions and relaxations), *essential tremor*, and sleep disorders such as *insomnia* (inadequate sleep), *hypersomnia* (excessive sleep), and sleep-related breathing problems (*sleep apnea* and *obstructive sleep apnea*). Beyond mental and neurological disorders, the robust enrichment of phenotypes also encompasses 7 *endocrine/metabolic* phenotypes, such as *vitamin B-complex deficiencies*, *hypothyroidism* and *hypopotassemia*; 6 *circulatory system* disorders, including *essential hypertension*, *cerebral atherosclerosis,* and *cerebral ischemia*; 5 genitourinary disorders including *urinary incontinence*; 4 musculoskeletal disorders such as *osteoarthrosis* and *osteoporosis*; 3 sense organ problems including *psychophysical visual disturbances*; and 3 digestive problems such as *blood in stool* and *hemorrhage of rectum and anus*.

**Table 2.**
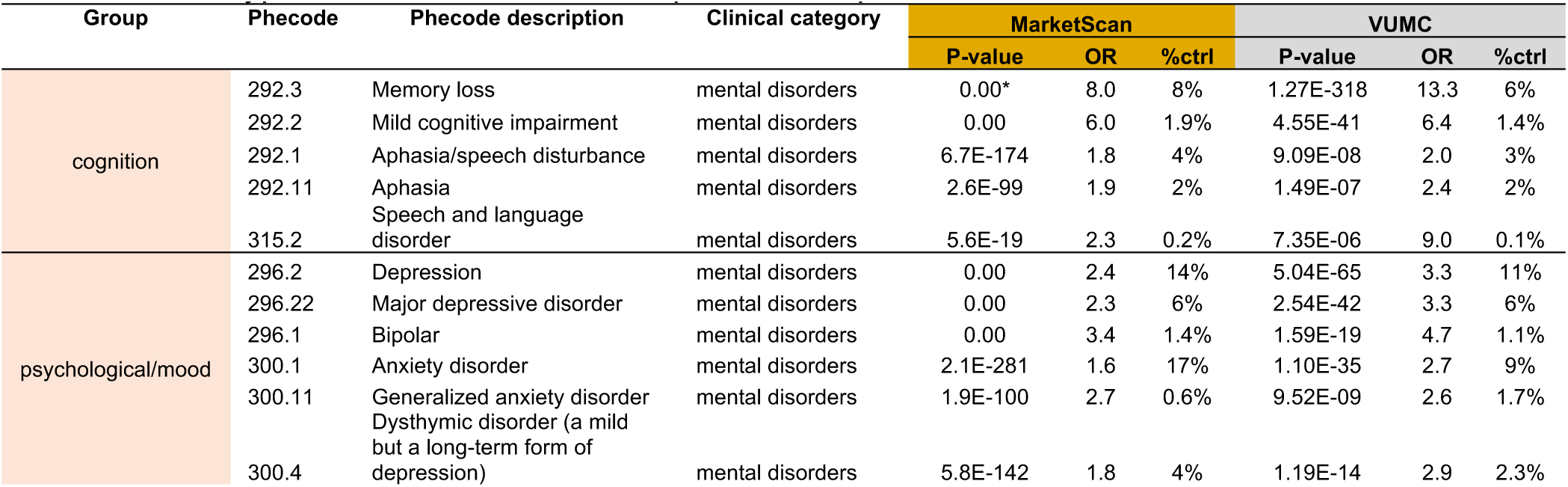
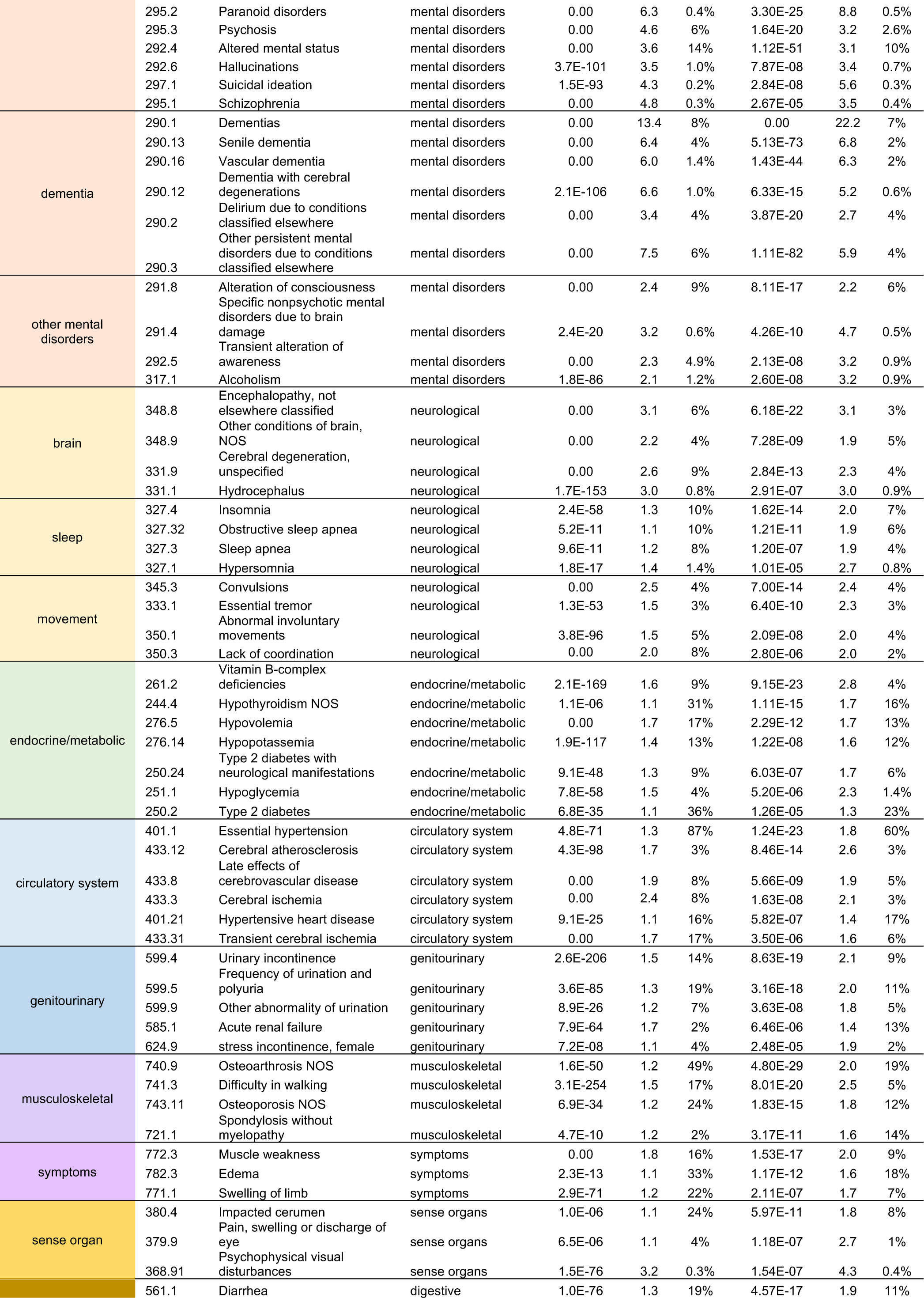
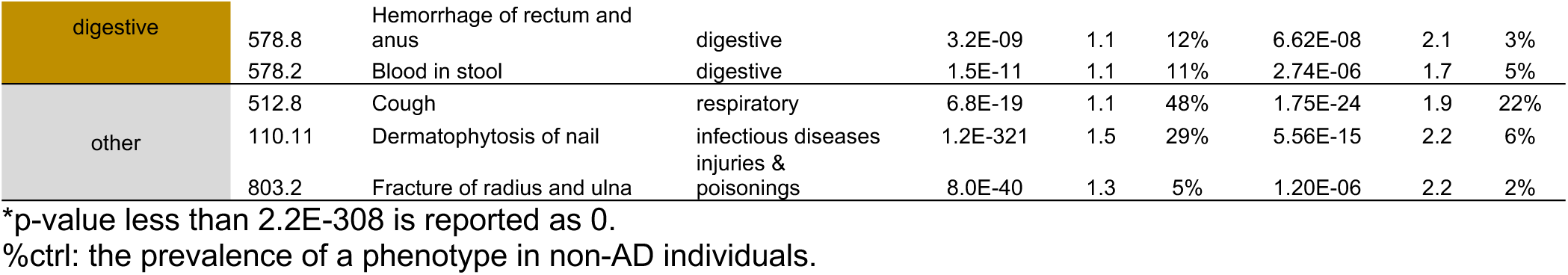
Phenotypes associated with subsequent development of AD in both EHR databases.

### A portion of the enriched phenotypes are genetically linked to AD

Having identified the phenotypes that are reliably detected in two independent large-scale EHR databases and are enriched in individuals at risk for future AD, we are now investigating the genetic underpinnings of these phenotypes. Phenotypes that share genetic susceptibility with AD and precede the clinical onset of AD may serve as early biomarkers before the irreversible cognition lapses. We compiled AD risk loci from the latest GWAS of AD and related dementia (ADRD)^16^ and constructed an instrumental variable by aggregating the independent ADRD risk alleles weighted by their effect size. The resulting polygenic risk score, PRS_AD_, consists of 78 independent signals including the APOE locus (**Suppl. Table S1**). Using the genotype data of European ancestry from BioVU (n=54,594), we calculated the PRS_AD_ for each person and performed PheWAS analysis across 1,493 phecodes. This analysis revealed 17 significant associations (at Bonferroni correction P<0.05/1493=3.3 × 10^−5^), including 5 phenotypes inversely associated with PRS_AD_. *Alzheimer’s disease* (*P*=7.3×10^−42^), *memory loss* (*P*=2.5×10^−20^), and *mild cognitive impairment* (*P*=5.2×10^−15^) are positively associated with PRS_AD_. *Dementias* (*P*=6.5×10^−50^) and subtypes such as *delirium dementia and amnestic and other cognitive disorder* (*P*=3.1×10^−32^), *senile dementia* (*P*=1.6×10^−7^), and *vascular dementia* (*P*=2.0×10^−7^) are also among the significant and positive associations. Other positive associations include *disorders of lipoid metabolism* (*P*=5.6×10^−8^), *hyperlipidemia* (*P*=7.4×10^−8^), *hypercholesterolemia* (*P*=1.1×10^−7^), and *neurological disorders* (*P*=1.5×10^−6^). Focusing on the phenotypes positively associated with PRS_AD_, we identified 9 such phenotypes that are not only showed enrichment prior to AD diagnosis but also are genetically linked to AD (**Table 3**). Upon excluding the APOE e4 allele (i.e. rs429358_C) from the PRS_AD_ and repeating the association analysis, none of the previously significant associations retained their significant (although nominal significance was observed for several phenotypes, such as *Alzheimer’s disease* (*P*=0.0009), *dementias* (*P*=0.0002), and *senile dementia* (*P*=0.005)) (**Suppl. Table S6**). This finding suggests that the observed phenotypic associations with PRS_AD_ are largely attributable to the APOE locus.

**Table 3.**
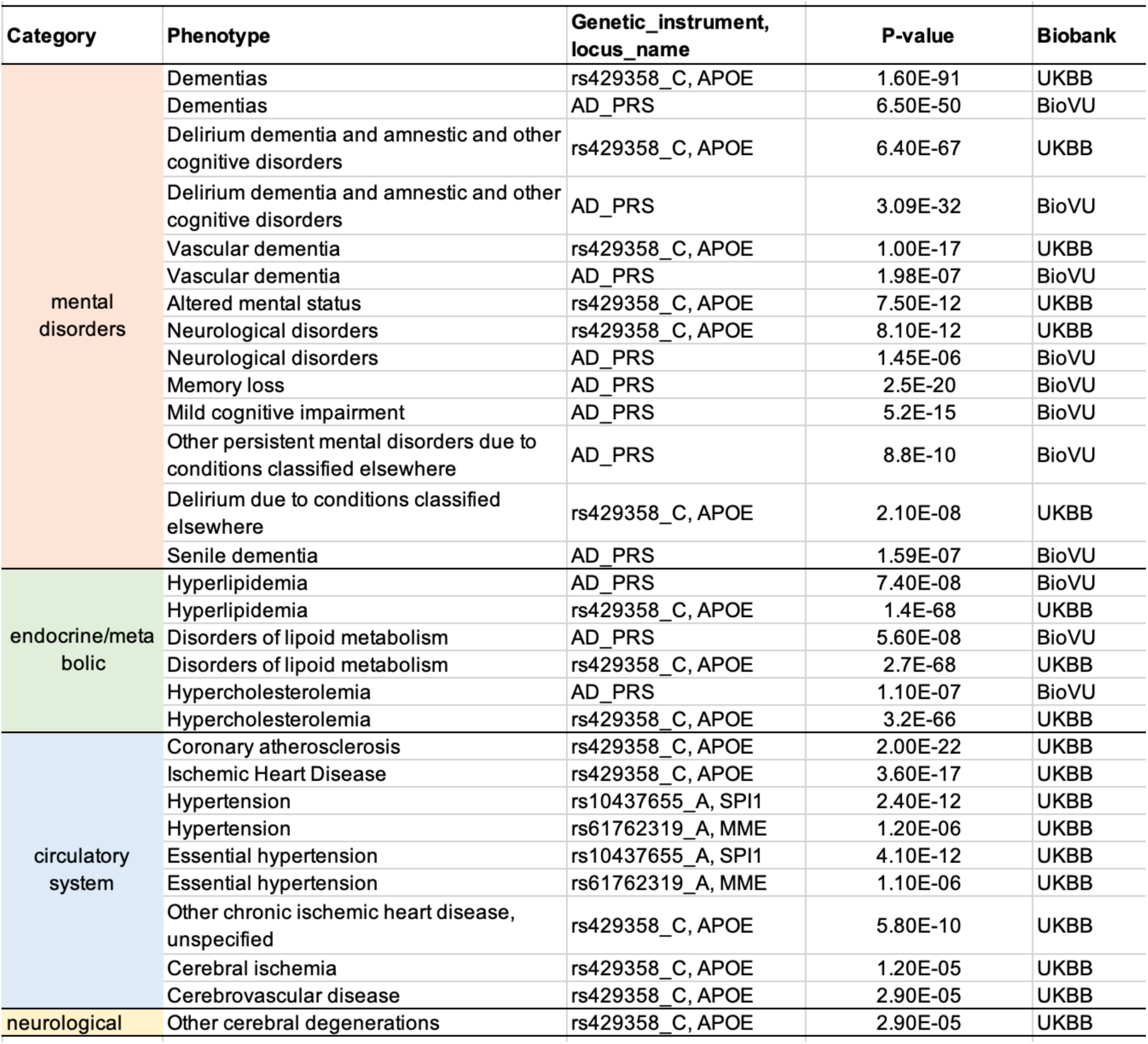
Phenotypes supported by AD genetics.

| Category | Phenotype | Genetic_instrument,<br>locus_name | P-value | Biobank |
| --- | --- | --- | --- | --- |
| mental disorders | Dementias | rs429358_C, APOE | 1.60E-91 | UKBB |
|  | Dementias | AD_PRS | 6.50E-50 | BioVU |
|  | Delirium dementia and amnestic and other cognitive disorders | rs429358_C, APOE | 6.40E-67 | UKBB |
|  | Delirium dementia and amnestic and other cognitive disorders | AD_PRS | 3.09E-32 | BioVU |
|  | Vascular dementia | rs429358_C, APOE | 1.00E-17 | UKBB |
|  | Vascular dementia | AD_PRS | 1.98E-07 | BioVU |
|  | Altered mental status | rs429358_C, APOE | 7.50E-12 | UKBB |
|  | Neurological disorders | rs429358_C, APOE | 8.10E-12 | UKBB |
|  | Neurological disorders | AD_PRS | 1.45E-06 | BioVU |
|  | Memory loss | AD_PRS | 2.5E-20 | BioVU |
|  | Mild cognitive impairment | AD_PRS | 5.2E-15 | BioVU |
|  | Other persistent mental disorders due to conditions classified elsewhere | AD_PRS | 8.8E-10 | BioVU |
|  | Delirium due to conditions classified elsewhere | rs429358_C, APOE | 2.10E-08 | UKBB |
|  | Senile dementia | AD_PRS | 1.59E-07 | BioVU |
| endocrine/metabolic | Hyperlipidemia | AD_PRS | 7.40E-08 | BioVU |
|  | Hyperlipidemia | rs429358_C, APOE | 1.4E-68 | UKBB |
|  | Disorders of lipid metabolism | AD_PRS | 5.60E-08 | BioVU |
|  | Disorders of lipid metabolism | rs429358_C, APOE | 2.7E-68 | UKBB |
|  | Hypercholesterolemia | AD_PRS | 1.10E-07 | BioVU |
|  | Hypercholesterolemia | rs429358_C, APOE | 3.2E-66 | UKBB |
| circulatory system | Coronary atherosclerosis | rs429358_C, APOE | 2.00E-22 | UKBB |
|  | Ischemic Heart Disease | rs429358_C, APOE | 3.60E-17 | UKBB |
|  | Hypertension | rs10437655_A, SPI1 | 2.40E-12 | UKBB |
|  | Hypertension | rs61762319_A, MME | 1.20E-06 | UKBB |
|  | Essential hypertension | rs10437655_A, SPI1 | 4.10E-12 | UKBB |
|  | Essential hypertension | rs61762319_A, MME | 1.10E-06 | UKBB |
|  | Other chronic ischemic heart disease, unspecified | rs429358_C, APOE | 5.80E-10 | UKBB |
|  | Cerebral ischemia | rs429358_C, APOE | 1.20E-05 | UKBB |
|  | Cerebrovascular disease | rs429358_C, APOE | 2.90E-05 | UKBB |
| neurological | Other cerebral degenerations | rs429358_C, APOE | 2.90E-05 | UKBB |

In addition to PRS_AD_, we also assessed the variant-level phenotypic associations using a larger biobank (UK Biobank) with 400,000 genotyped white British samples. We interrogated the phenome-wide associations and yielded 82 significant associations (P<0.05/1400=3.6×10^−5^) involving 17 AD risk loci (**Suppl. Table S7**). The AD risk allele at the APOE locus (rs429358_C) is positively associated with 17 phenotypes, recapitulating the phenotypes seen for the associations with PRS_AD_. These include *Alzheimer’s disease* (*P*=4.3×10^−66^), *dementias* (*P*=1.6×10^−91^), *vascular dementia* (*P*=1.0×10^−17^), *hyperlipidemia* (*P*=1.4×10^−68^), *disorders of lipoid metabolism* (*P*=2.7×10^−68^), and *hypercholesterolemia* (*P*=3.2×10^−66^). In addition to mental and endocrine/metabolic disorders, the APOE e4 risk allele was also associated with an increased risk of circulatory system disorders, such as *coronary atherosclerosis* (*P*=2.0×10^−22^), *ischemic heart disease* (*P*=3.6×10^−17^), *cerebral ischemia* (*P*=3.2×10^−5^), and *cerebrovascular disease* (*P*=2.9×10^−5^). *Hypertension* was associated with two other AD risk alleles, located at the SPI1 locus (rs10437655_A) and MME locus (rs61762319_A), respectively. In total, there are 22 unique phenotypes that are robustly associated with a higher risk of AD are supported by AD genetics, through either PRS or individual risk alleles, or both (**Table 3**).

### Gender effect

Given that women are at a higher risk of developing AD than men, we were curious to determine whether any of the phenotypic enrichment observed was more pronounced in one gender over the other. For this purpose, we stratified the MS samples by gender and re-estimated the enrichment for each gender. To ensure a fair comparison, we focused on general phenotypes by excluding male-only and female-only phenotypes (medical conditions involving sexual organs). Overall, of the phenotypes that reached nominal significance in both genders (n=286), no pronounced gender effect was detected: the resulting enrichment estimates were consistent between the genders (regression line: male=1.0334×female+0.0243 for the natural logarithm of the ORs, correlation=0.893) (**Suppl. Table S8**, **Suppl. Fig S1**). This indicates that our matching approach has effectively mitigated potential gender bias, and the resulting odds ratio estimates reflect a gender-independent enrichment.

### Sensitivity analysis

#### Co-occurrence of AD with other dementias and its impact on enrichment estimation

It is well documented that AD can present along with other dementia types ^26–29^. We confirmed this phenomenon of co-occurrence of AD and other dementias in EHRs. For example, in MarketScan, approximately 60% of individuals diagnosed with AD had previously been diagnosed with another type of dementia. To investigate whether such co-occurrence may have influenced the enrichment estimation, we conducted an analysis within the larger MS database. Specifically, we excluded AD cases that had any prior diagnosis of other dementia (*dementias*, *senile dementia*, *vascular dementia*, *dementia with cerebral degeneration*, *delirium dementia and amnestic, and other cognitive disorders*) (n=20,153) (**Suppl. Table S9**). With exclusion of these AD cases, their matched controls were also excluded from analysis (n=251,030) (**Suppl. Table S9**). The re-estimated odds ratios from this subset of data revealed a reduction in associated phenotypes (after Bonferroni correction, *P*<2.9×10^−5^) (**Suppl. Fig. S2A-B**). Mental disorders are markedly reduced to half or less of their original values, for example, *delirium due to conditions classified elsewhere* (new OR=2.0 vs. original 4.0), *developmental delays and disorders* (OR=2.1 vs. original 5.4), *paranoid disorders* (OR=3.5 vs. original 7.4), *psychosis* (OR=2.6 vs. original 5.1), *conduct disorders* (OR=3.1 vs. original 7.4), and *suicidal ideation* (OR=2.3 vs. original 4.6). In contrast, the hallmark manifestations of AD had little changes in the resulting estimates: *memory loss* (OR=9.1 vs. original 9.5) and *mild cognition impairment* (OR=6.3 vs. original 7.3). This suggests that the inclusion or exclusion of AD cases with comorbidity of other dementias did not significantly affect the estimates of AD-specific manifestations, while the psychological symptoms that were heavily affected are more generic to all dementia types. Finally, we note that there was no difference in the prevalence of medical conditions observed in the control samples between the original and subset datasets (**Suppl. Fig. S2C**), which reassures us that the observed attenuation in estimates is not due to the comparison of the full versus subset control samples.

## DISCUSSION

EHR data, a rapidly expanding yet underutilized resources, holds significant potential for AD research. With access to longitudinal EHRs from over 150 million individuals, we possess the statistical power for a systematic interrogation of medical conditions that are more likely to occur in individuals who later develop AD. By comparing the documented prevalence of medical conditions between AD patients in later life and age– and gender-matched controls over an equivalent observation period, we identified and validated 73 medical phenotypes that showed enrichment prior to AD diagnosis in both the MarketScan and VUMC databases. Among these phenotypes, mental disorders comprise one-third of the total, neurological disorders alone comprise one-sixth, endocrine/metabolic and circulatory disorders together make up another one-sixth, and the remaining third is represented by nine distinct clinical categories. Our genetic analysis further revealed connections between a small portion of these phenotypes and AD genetics, showing associations with specific AD risk variants or PRS.

Our study confirms hypertension and hypercholesterolemia as risk factors for the development of late-life AD. Without cure for dementia (the current FDA-approved amyloid-based treatment for AD is intended for patients with early/mild Alzheimer’s disease), risk reduction and prevention are of paramount importance. Modifying these risk factors through the adoption of healthier lifestyles or the use of anti-hypertensive or lipid-lowering medications that are more brain-healthy can be effective measures for risk reduction. Ongoing clinical trials for AD and related dementia are investigating several anti-hypertensives (telmisartan, losartan, amlodipine and perindopril) and lipid-lowering medication (atorvastatin)^30^. Findings from these clinical trials will provide evidence on the protective effects of these medications against dementia and will guide the selection of the most appropriate interventions.

Previous studies have noted the absence of a significant association between overall AD genetics and neuropsychiatric diseases. A study of genetic sharing among 25 brain disorders revealed extremely limited extent of genetic correlation between AD and psychiatric disorders (e.g., anxiety and major depressive disorder) as well as neurodegenerative disease (e.g., Parkinson’s disease) ^31^. In a GWAS of AD involving 71,880 clinically diagnosed AD cases and proxies, along with 383,378 controls, the genetic correlation between AD and depression barely reached significance ^32^. Despite the limited genetic sharing at the global scale between AD and psychiatric diseases, at individual genetic loci, some are indeed found to be shared between AD and psychiatric disorders ^33,34^. Local genetic correlation analysis from a recent study ^35^ further revealed four GWAS loci (e.g. the TMEM106B locus at chr7 or rs13237518) that are shared between depression and AD, five loci shared between schizophrenia and AD, and two loci shared between bipolar and AD. Therefore, among individuals with depression, bipolar, or schizophrenia, those who carry AD-causing variants may be more susceptible to developing AD than others.

An intriguing observation is that neoplasms consistently emerge as depleted phenotypes in both EHR datasets. This observation is consistent with the inverse relationship between cancer diagnosis and Alzheimer’s disease reported recently, a trend supported by accumulating evidence ^36–40^. It has been hypothesized that age-related alterations in metabolism and energy balance may concurrently increase the risk of AD while lowering the risk of cancer, or vice versa ^37,41,42^; however, the exact underlying mechanisms remain elusive. The situation is further complicated by the fact that cognitive impairment, often referred to as “brain fog,” has been reported as an adverse effect associated with cancer treatment ^43^.

The study has several limitations. The use of the first recorded diagnosis code of AD in the EHRs may not faithfully capture the actual date of AD onset, as the actual emergence of AD could have been earlier. Nonetheless, we observed in both EHR datasets that 80% of the AD patients received their first diagnosis code of AD at or above the age of 75, which aligns with the typical age distribution for late-onset AD. Second, we only used diagnosis codes (ICD) to identify case/control status of AD, which may result in under-detection of AD cases. The integration of medication and clinical notes may enhance the detection of AD cases ^44^. In both EHRs datasets, the identified AD patients comprise approximately 0.30% of the total population, a figure lower than the national prevalence of AD cases, which is about 2% when considering the estimated 6.7 million Americans living with AD out of a total US population of 335 million as of 2023. The under-detection of AD cases may result in the inclusion of “unrecognized AD cases” within the control group, leading to a conservative attenuation in the enrichment estimation between AD and controls. Consequently, the current list of enriched phenotypes is likely to remain robust, if not more pronounced.

## CONCLUSION

In conclusion, our systematic interrogation of two independent large-scale EHR databases with 153 million individuals demonstrates the feasibility of utilizing the growing resources of longitudinal EHRs to substantially expand the list of health conditions for association with subsequent development of AD. By tracking and comparing the documented medical conditions over an equivalent observation period prior to AD diagnosis, we discovered and replicated more than 70 medical phenotypes that occurred more likely in those who later received AD diagnosis compared to their age– and gender-matched counterparts. Our findings reveal that the associated phenotypes are dominated by neuropsychological, circulatory and endocrine/metabolic disorders. Furthermore, about a quarter of the phenotypes demonstrate sharing with AD genetics. Together, our findings highlight potential opportunities of therapeutics and interventions to reduce AD risk.

## Declarations

### Ethics approval and consent to participate

All human subjects used in this study have provided informed consent by the previous studies. Consent was not necessary for this study.

### Consent for publication

All authors approved the final version of the manuscript and consent for publication.

### Availability of data and material

Information on how to collaborate and the BioVU data-sharing policy can be found at https://victr.vumc.org/how-to-use-biovu/. Access to the variant-level phenome-wide association results based on UK Biobank data are available at https://pheweb.sph.umich.edu. PheWAS mapping can be find at PheWAS sources (https://phewascatalog.org/phewas/).

### Competing interests

The authors declare no competing interests for this work.

### Author contributions

X.Z. and N.J.C. conceived and designed the study. G.J. and A.R. acquired and pre-processed the MarketScan data; Y.Z. and K.C. extracted and pre-processed the VUMC EHR data. N.J.C., X.Z. and B.L. obtained access to the BioVU data. X.Z. conducted formal analyses, prepared the Figures/Tables, and drafted the manuscript. B.L. acquired the funding. All authors participated in the interpretation of the results, reviewed and/or edited, and approved the submitted version of the manuscript.

## Data Availability

All data produced in the present work are contained in the manuscript.

https://victr.vumc.org/how-to-use-biovu/

https://pheweb.sph.umich.edu

https://phewascatalog.org/phewas/

## Abbreviations

AD: Alzheimer’s disease
ADRD: Alzheimer’s disease and related dementia
BioVU: Biobank of Vanderbilt University
EHR: Electronic health records
GWAS: Genome-wide association study
ICD9: International classification of disease version 9
ICD10: International classification of disease version 10
OR: Odds ratio
PCA: Principal component analysis
PheWAS: Phenome-wide association study
PRS: Polygenic risk score
SD: Synthetic Derivative
VUMC: Vanderbilt University Medical Center

## Acknowledgements

X.Z is supported by NIA grant R01AG069900; Y.Z is supported by NHGRI grants U54HG012510 and RM1HG009034; A.R. is supported by NIMH grant R01MH137646 and J&J contract C2021013133; B.L. is supported by NIA grants R01AG069900, R01AG089717 and R01AG065611.

The dataset used for part of the genetic-association analyses were obtained from Vanderbilt University Medical Center’s BioVU biorepository, which is supported by institutional funding, private agencies and federal grants, including the NIH-funded S10RR025141 instrumentation award and Clinical and Translational Science Award grants UL1TR002243, UL1TR000445 and UL1RR024975; genomic data are also supported by investigator-led projects that include U01HG004798, R01NS032830, RC2GM092618, P50GM115305, U01HG006378, U19HL065962 and R01HD074711, as well as the additional funding sources listed at https://victr.vanderbilt.edu/pub/biovu/. The funders had no role in study design, data collection, analysis, decision to publish, or preparation of the manuscript. We gratefully acknowledge the openly available summary statistics from the Michigan University Pheweb resource.

## Notes

### Competing Interest Statement

The authors have declared no competing interest.

## References

1 Burns, A. & Iliffe, S. Alzheimer’s disease. BMJ 338, b158 (2009). 10.1136/bmj.b158

2 Sevigny, J. et al. The antibody aducanumab reduces Abeta plaques in Alzheimer’s disease. Nature 537, 50–56 (2016). 10.1038/nature19323

3 Sims, J. R. et al. Donanemab in Early Symptomatic Alzheimer Disease: The TRAILBLAZER-ALZ 2 Randomized Clinical Trial. JAMA 330, 512–527 (2023). 10.1001/jama.2023.13239

4 van Dyck, C. H. et al. Lecanemab in Early Alzheimer’s Disease. N Engl J Med 388, 9–21 (2023). 10.1056/NEJMoa2212948

5 Kivipelto, M. et al. Midlife vascular risk factors and Alzheimer’s disease in later life: longitudinal, population based study. BMJ 322, 1447–1451 (2001). 10.1136/bmj.322.7300.1447

6 Kivipelto, M. et al. Risk score for the prediction of dementia risk in 20 years among middle aged people: a longitudinal, population-based study. Lancet Neurol 5, 735–741 (2006). 10.1016/S1474-4422(06)70537-3

7 Alber, J. et al. Developing retinal biomarkers for the earliest stages of Alzheimer’s disease: What we know, what we don’t, and how to move forward. Alzheimers Dement 16, 229–243 (2020). 10.1002/alz.12006

8 Bayer, A. U., Ferrari, F. & Erb, C. High occurrence rate of glaucoma among patients with Alzheimer’s disease. Eur Neurol 47, 165–168 (2002). 10.1159/000047976

9 Green, R. C. et al. Depression as a risk factor for Alzheimer disease: the MIRAGE Study. Arch Neurol 60, 753–759 (2003). 10.1001/archneur.60.5.753

10 Holmquist, S., Nordstrom, A. & Nordstrom, P. The association of depression with subsequent dementia diagnosis: A Swedish nationwide cohort study from 1964 to 2016. PLoS Med 17, e1003016 (2020). 10.1371/journal.pmed.1003016

11 Zissimopoulos, J., Crimmins, E. & St Clair, P. The Value of Delaying Alzheimer’s Disease Onset. Forum Health Econ Policy 18, 25–39 (2014). 10.1515/fhep-2014-0013

12 Ponjoan, A. et al. How well can electronic health records from primary care identify Alzheimer’s disease cases? Clin Epidemiol 11, 509–518 (2019). 10.2147/CLEP.S206770

13 Wei, W. Q. et al. Evaluating phecodes, clinical classification software, and ICD-9-CM codes for phenome-wide association studies in the electronic health record. PLoS One 12, e0175508 (2017). 10.1371/journal.pone.0175508

14 Wilkinson, T. et al. Identifying dementia cases with routinely collected health data: A systematic review. Alzheimers Dement 14, 1038–1051 (2018). 10.1016/j.jalz.2018.02.016

15 Denny, J. C. et al. PheWAS: demonstrating the feasibility of a phenome-wide scan to discover gene-disease associations. Bioinformatics 26, 1205–1210 (2010). 10.1093/bioinformatics/btq126

16 Bellenguez, C. et al. New insights into the genetic etiology of Alzheimer’s disease and related dementias. Nat Genet 54, 412–436 (2022). 10.1038/s41588-022-01024-z

17 Jia, G. et al. Estimating heritability and genetic correlations from large health datasets in the absence of genetic data. Nat Commun 10, 5508 (2019). 10.1038/s41467-019-13455-0

18 Putting Research Data into Your Hands with the MarketScan Databases 2016. *Available at:* http://truvenhealth.com/markets/life-sciences/products/data-tools/marketscan-databases *[Accessed 2020 Feb 6]*.

19 Roden, D. M. et al. Development of a large-scale de-identified DNA biobank to enable personalized medicine. Clin Pharmacol Ther 84, 362–369 (2008). 10.1038/clpt.2008.89

20 Do, R. et al. Common variants associated with plasma triglycerides and risk for coronary artery disease. Nat Genet 45, 1345–1352 (2013). 10.1038/ng.2795

21 McCarthy, S. et al. A reference panel of 64,976 haplotypes for genotype imputation. Nat Genet 48, 1279–1283 (2016). 10.1038/ng.3643

22 Howie, B. N., Donnelly, P. & Marchini, J. A flexible and accurate genotype imputation method for the next generation of genome-wide association studies. PLoS Genet 5, e1000529 (2009). 10.1371/journal.pgen.1000529

23 Carroll, R. J., Bastarache, L. & Denny, J. C. R PheWAS: data analysis and plotting tools for phenome-wide association studies in the R environment. Bioinformatics 30, 2375–2376 (2014). 10.1093/bioinformatics/btu197

24 Taliun, D. et al. Sequencing of 53,831 diverse genomes from the NHLBI TOPMed Program. Nature 590, 290–299 (2021). 10.1038/s41586-021-03205-y

25 Zhou, W. et al. Efficiently controlling for case-control imbalance and sample relatedness in large-scale genetic association studies. Nat Genet 50, 1335–1341 (2018). 10.1038/s41588-018-0184-y

26 Kovacs, G. G. et al. Non-Alzheimer neurodegenerative pathologies and their combinations are more frequent than commonly believed in the elderly brain: a community-based autopsy series. Acta Neuropathol 126, 365–384 (2013). 10.1007/s00401-013-1157-y

27 Rahimi, J. & Kovacs, G. G. Prevalence of mixed pathologies in the aging brain. Alzheimers Res Ther 6, 82 (2014). 10.1186/s13195-014-0082-1

28 Robinson, J. L. et al. The development and convergence of co-pathologies in Alzheimer’s disease. Brain 144, 953–962 (2021). 10.1093/brain/awaa438

29 Robinson, J. L. et al. Pathological combinations in neurodegenerative disease are heterogeneous and disease-associated. Brain 146, 2557–2569 (2023). 10.1093/brain/awad059

30 Cummings, J. et al. [Not Available]. Alzheimers Dement (N Y) 10, e12465 (2024). 10.1002/trc2.12465

31 Brainstorm, C. et al. Analysis of shared heritability in common disorders of the brain. Science 360 (2018). 10.1126/science.aap8757

32 Jansen, I. E. et al. Genome-wide meta-analysis identifies new loci and functional pathways influencing Alzheimer’s disease risk. Nat Genet 51, 404–413 (2019). 10.1038/s41588-018-0311-9

33 Harerimana, N. V. et al. Genetic Evidence Supporting a Causal Role of Depression in Alzheimer’s Disease. Biol Psychiatry 92, 25–33 (2022). 10.1016/j.biopsych.2021.11.025

34 Wingo, T. S. et al. Shared mechanisms across the major psychiatric and neurodegenerative diseases. Nat Commun 13, 4314 (2022). 10.1038/s41467-022-31873-5

35 Ajneesh Kumar, N. R. R., Michael L. Cuccaro, Gary Beecham, & Edward D. Huey, C. R. Genetic correlation analysis identifies genetic loci shared between Alzheimer’s disease and primary psychiatric disorders. (https://alz.confex.com/alz/2024/meetingapp.cgi/Paper/93421, Alzheimer’s Association International Conference2024).

36 Aramillo Irizar, P., et al. Transcriptomic alterations during ageing reflect the shift from cancer to degenerative diseases in the elderly. Nat Commun 9, 327 (2018). 10.1038/s41467-017-02395-2

37 Driver, J. A. Inverse association between cancer and neurodegenerative disease: review of the epidemiologic and biological evidence. Biogerontology 15, 547–557 (2014). 10.1007/s10522-014-9523-2

38 Driver, J. A. et al. Inverse association between cancer and Alzheimer’s disease: results from the Framingham Heart Study. BMJ 344, e1442 (2012). 10.1136/bmj.e1442

39 Ospina-Romero, M. et al. Rate of Memory Change Before and After Cancer Diagnosis. JAMA Netw Open 2, e196160 (2019). 10.1001/jamanetworkopen.2019.6160

40 Roe, C. M. et al. Cancer linked to Alzheimer disease but not vascular dementia. Neurology 74, 106–112 (2010). 10.1212/WNL.0b013e3181c91873

41 Demetrius, L. A. & Driver, J. A. Preventing Alzheimer’s disease by means of natural selection. J R Soc Interface 12, 20140919 (2015). 10.1098/rsif.2014.0919

42 Okereke, O. I. & Meadows, M. E. More Evidence of an Inverse Association Between Cancer and Alzheimer Disease. JAMA Netw Open 2, e196167 (2019). 10.1001/jamanetworkopen.2019.6167

43 Ahles, T. A. Brain vulnerability to chemotherapy toxicities. Psychooncology 21, 1141–1148 (2012). 10.1002/pon.3196

44 Wei, W. Q. et al. Combining billing codes, clinical notes, and medications from electronic health records provides superior phenotyping performance. J Am Med Inform Assoc 23, e20–27 (2016). 10.1093/jamia/ocv130

