## Supplementary_Figures_S1-S2 for "Longitudinal Analysis of Electronic Health Records Reveals Medical Conditions Associated with Subsequent Alzheimer’s Disease Development"

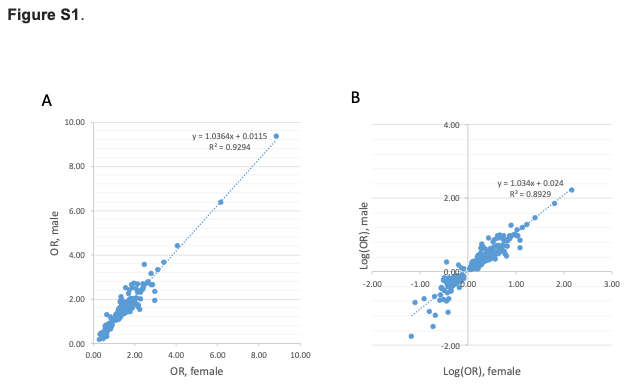
**Figure S1**. Gender difference is minimal for phenotypes whose enrichment are detected in both genders. (A) enrichment odds ratios (OR) and fitted line; (B) logarithm of enrichment odds ratios and fitted line.

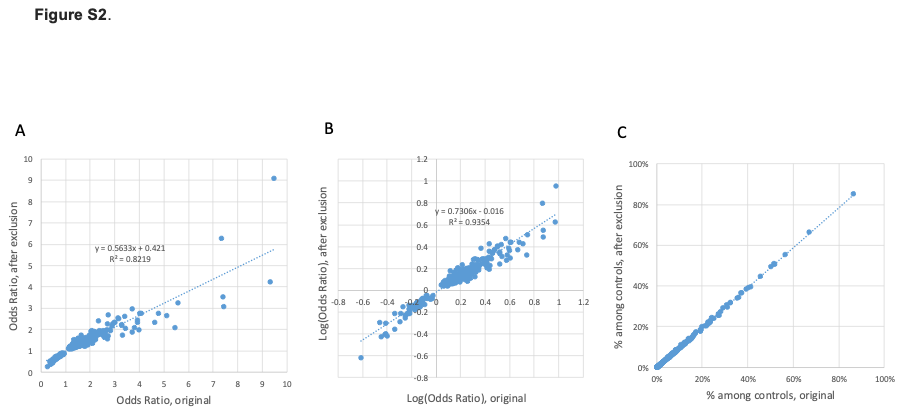

**Figure S2**. Enrichment estimates with and without exclusion of AD cases with previous diagnosis of other dementias and the corresponding matched controls. (A) enrichment odds ratios and fitted line; (B) logarithm of enrichment odds ratios and fitted line; (C) prevalence (%) of medical phenotypes among the reference populations (i.e. non-AD controls).
